# A More-Than-Human Approach to Designing for Mental Health: Remixing Prototypes for the Contexts of Complex Healthcare Infrastructures

**DOI:** 10.64898/2026.06.10.26355412

**Authors:** Vincent Allen, Karolina Stasiak, Danielle Lottridge

**Affiliations:** Department of Psychological Medicine, The University of Auckland, Private Bag 92019, Auckland 1142, New Zealand; Department of Computer Science, The University of Auckland, Private Bag 92019, Auckland 1142, New Zealand

**Keywords:** digital mental health, more-than-human design, contextual inquiry, healthcare infrastructure, iterative prototyping

## Abstract

Digital mental health tools (DMHTs) often fail to be successfully implemented in clinical settings. While user- and human-centred design frameworks are frequently proposed for developing effective tools, they are insufficient to address the sociotechnical complexity of healthcare environments. This paper addresses this limitation by detailing the application of a more-than-human design framework to incorporate wider contextual factors into design decisions.

To demonstrate the application of this more-than-human design framework, we present a case study showcasing the design of one specific feature within a DMHT intended to support Health Improvement Practitioners (HIPs) in New Zealand’s Integrated Primary Mental Health and Addictions (IPMHA) service. Our process blends usage-context storyboards with interface prototypes, using think-aloud interviews to test the contextual fit of our prototypes.

The initial design concept failed due to contextual factors such as inconsistent wait times and the administrative burden on clients and clinic staff. This led to a pivot to a more context-appropriate, practitioner-focused, in-session concept for digital psychometric administration and automated scoring. This case study demonstrates that for DMHTs to be viable within complex healthcare environments, design must focus on more than the needs of a single user, incorporating multiple stakeholders and contextual variables across the wider service-delivery context.

## 1. Introduction

### 1.1 Using Digital Technology to Enhance Mental Health Service Delivery

Worldwide, nearly one billion people are affected by mental health conditions, with most people unable to access effective mental health care (World Health Organization [WHO], 2022). This treatment gap is a result of chronic underinvestment in mental health services, leading to systemic barriers such as stigma around service access, workforce shortages, and financial and geographical limitations (WHO, 2022). In response to this global crisis, health systems and innovators are increasingly turning to digital technology to help bridge the treatment gap. Digital mental health tools (DMHTs) are proposed as a scalable solution with the potential to overcome many traditional barriers by offering services that can be more accessible and affordable than conventional care. By leveraging technology, these tools aim to bypass geographical limitations, reduce financial burdens, and mitigate the stigma that often prevents individuals from seeking help (WHO, 2022).

While the term ’digital mental health tool’ typically brings to mind standalone self-guided smartphone applications, such as popular wellness apps like Calm (Calm.com, Inc., 2025) or Headspace (Headspace, Inc., 2025), the field is far broader. DMHTs encompass a range of technologies and delivery formats. These include guided interventions such as Thought-Spot (Thought-Spot Limited, 2025) and DBT Coach (Swasth Inc., 2025), where practitioners offer asynchronous support, as well as fully blended-care models such as Lyra Care Therapy (Espel-Huynh et al., 2024) where the digital tool is used as a core component of a formal treatment plan alongside face-to-face therapy. Although standalone self-help DMHTs are the most prevalent (EPHA, 2024) and have demonstrated modest reductions in depression and anxiety symptoms (Linardon et al., 2024; Seegan et al., 2023), they may be less appropriate for treating more severe or complex mental health conditions (Philippe et al., 2022).

A critical determinant of a DMHT’s clinical effectiveness is the inclusion of human guidance (Moshe et al., 2021; Nunes-Zlotkowski et al., 2024; Philippe et al., 2022). This necessity provides a compelling argument for integrating digital tools into existing clinical workflows, moving beyond standalone, self-guided applications toward guided and blended-care models capable of treating complex and severe conditions. Recent meta-analyses have demonstrated that the professional qualification of the individual providing this human guidance does not impact DMHT efficacy (Leung et al., 2022; Moshe et al., 2021). This finding establishes a strong, evidence-based foundation for utilising non-specialist practitioners or allied support staff to guide patients through integrated DMHTs, directly addressing systemic access limitations driven by specialist workforce shortages.

### 1.2 Digital Technology to Support Integrated Primary Mental Health Services

The global pattern of poor mental health service access (WHO, 2022) is reflected in New Zealand, with demand for mental health services growing (Ministry of Health, 2020; Ministry of Health, 2024; Statistics New Zealand, 2021) and an under-resourced public health system struggling to meet the escalating need (Health and Disability Commissioner, 2020).

Over two decades ago, calls for global reform highlighted the need for new models of care, particularly advocating for the integration of mental health services into primary care clinics to improve access and reduce stigma (WHO, 2001). Despite this long-standing recommendation, progress has been slow. While some nations have invested in the development of integrated primary services, many continue to rely on conventional, specialist-driven approaches that lack the capacity and scalability to address the population’s needs (WHO, 2022).

New Zealand is one of the nations that has made significant investments in this area with the development of the Integrated Primary Mental Health and Addictions (IPMHA) model (Ministry of Health, 2022). This model was adapted from the widely used Primary Care Behavioural Health (PCBH) model (Robinson & Reiter, 2015) to address the specific challenges of the New Zealand primary care service-delivery context. First piloted in 2017 (Appleton-Dyer, 2018), the model embeds behavioural health clinicians, known as Health Improvement Practitioners (HIPs), directly into general practice teams (Ministry of Health, 2022), using Focused Acceptance and Commitment Therapy (fACT) to deliver brief single-session interventions (Strosahl et al., 2012).

The IPMHA model was designed for rapid nationwide scaling (Ministry of Health, 2022). By July 2024, these services were available to over 70% of the enrolled primary care population (Te Whatu Ora, 2025). A core feature of the IPMHA model enabling this expansion was a workforce development strategy designed to overcome the issue of insufficient specialist mental health service providers in New Zealand. In contrast to the PCBH model which requires practitioners to have specialised postgraduate degrees, the IPMHA model recruits registered health professionals from diverse backgrounds, such as nursing, social work, and occupational therapy, and upskills them through a brief one- to two-week training programme (Te Pou, 2024). While this has helped to address workforce shortages, the brief nature of the training and non-specialist workforce raises concerns about service quality and consistency (Ministry of Health, 2022).

Evaluations of the IPMHA model have shown promising results, with a 2018 pilot demonstrating improved service access (Appleton-Dyer, 2018). A subsequent, larger evaluation by New Zealand’s Ministry of Health (Ministry of Health, 2022) confirmed these benefits, highlighting reduced wait times, decreased stigma, and positive patient experiences. However, this report also identified significant implementation challenges, including inconsistencies across regions, difficulties in maintaining model fidelity, practitioner training challenges, and the need for ongoing workforce development to meet service demand (Ministry of Health, 2022). Building on the findings of this evaluation report, a recent study looking at the experiences of HIPs within the IPMHA model (Allen et al., 2024) identified a number of service-delivery challenges. These included: the negative impact of overly brief sessions, insufficient training, a lack of culturally appropriate resources, high workloads, administrative inefficiencies, difficult referrals, and inadequate client follow-up. Despite the successes of the model, a substantial treatment gap remains. While 32.1% of New Zealand adults report experiencing moderate to severe psychological distress, recent data shows that over a one-year period only 2.9% of adults who visited a primary care clinic engaged with integrated mental health service providers (Ministry of Health, 2024). This gap between population-level need and service uptake demonstrates that while making services available within primary care has helped to improve access, population needs still outpace uptake.

The IPMHA model represents a significant advancement in improving access to mental health support in New Zealand, yet service-delivery challenges persist and the model struggles to address the full scale of the service demand (Ministry of Health, 2024). To enhance the scalability and reach of integrated care approaches like the IPMHA model, innovative solutions are required that do not rely on the limited capacity of the healthcare workforce. Digital technology has been identified as a promising approach for augmenting existing service models (Ministry of Health, 2022; WHO, 2022), offering a potential pathway to strengthen the effectiveness and reach of the IPMHA model in the New Zealand primary care context.

### 1.3 Poor Contextual Fit Leads to Low Practitioner Digital Adoption

For a guided or blended-care DMHT to be successfully implemented within an existing service-delivery context, it must be used and endorsed by the practitioner delivering the service, making practitioner adoption an essential prerequisite for success (Mohr et al., 2025). However, low practitioner adoption remains one of the primary barriers to effective implementation of DMHTs into existing services (Berardi et al., 2024; Torous et al., 2025). A synthesis of several systematic reviews reveals a set of common barriers to clinician adoption of digital health tools across various healthcare service-delivery contexts (Berardi et al., 2024; Borges do Nascimento et al., 2023; Gagnon et al., 2016; Jacob et al., 2020; Zakerabasali et al., 2021). The most prevalent barriers consistently reported across different healthcare services included: 1) Workload and workflow disruption, 2) inadequate training and support, 3) concerns about privacy and security risks, and for mental health services specifically 4) the potential impact on the therapeutic relationship.

The scope of these barriers suggests that practitioner adoption is not merely a technical problem, but a complex sociotechnical one. A DMHT’s success is determined less by its technological sophistication and more by its contextual fit: how well it aligns with the social, cultural, and organisational realities of the clinical environment (Jacob et al., 2020). While the literature comprehensively identifies common adoption barriers and broadly recommends the use of human-centred and co-design frameworks to overcome them, there is a significant gap in the literature concerning practical evidence-based strategies for the proactive design and implementation of DMHTs into complex healthcare environments (Mohr et al., 2025).

The gap in evidence-based design strategy for DMHTs is exacerbated by the commercial nature of research and development in this field. Since many tools are developed through private partnerships between technology companies and healthcare providers, their design processes and findings are rarely shared publicly (Mohr et al., 2025). This practice traps valuable knowledge within individual organisations, creating knowledge silos. This lack of transparency undermines efforts to build a cumulative evidence base, hindering the creation of effective DMHTs that can be integrated into healthcare systems. This environment poses a significant challenge for academic researchers who often lack access to in-house software development or design expertise (Mohr et al., 2025). This project will address the identified gap by describing the process of applying a context-sensitive ‘more-than-human’ design framework (Coskun et al., 2022; Eriksson et al., 2024) to the ideation and prototyping development stages for a DMHT tailored to the needs of the IPMHA service-delivery context.

### 1.4 Embracing Complexity: Adopting a Context-Sensitive More-than-Human Design Framework

The foundational design framework chosen for this project was the five-stage Design Thinking model (*see Figure 1*), a user-centred, solution-oriented, iterative problem-solving approach (Brown, 2008; Interaction Design Foundation, 2016). It emphasises the importance of collaborative design with stakeholders to clearly understand user needs, generate innovative solutions, and test them through an ongoing iterative prototyping process.

**Figure 1.**
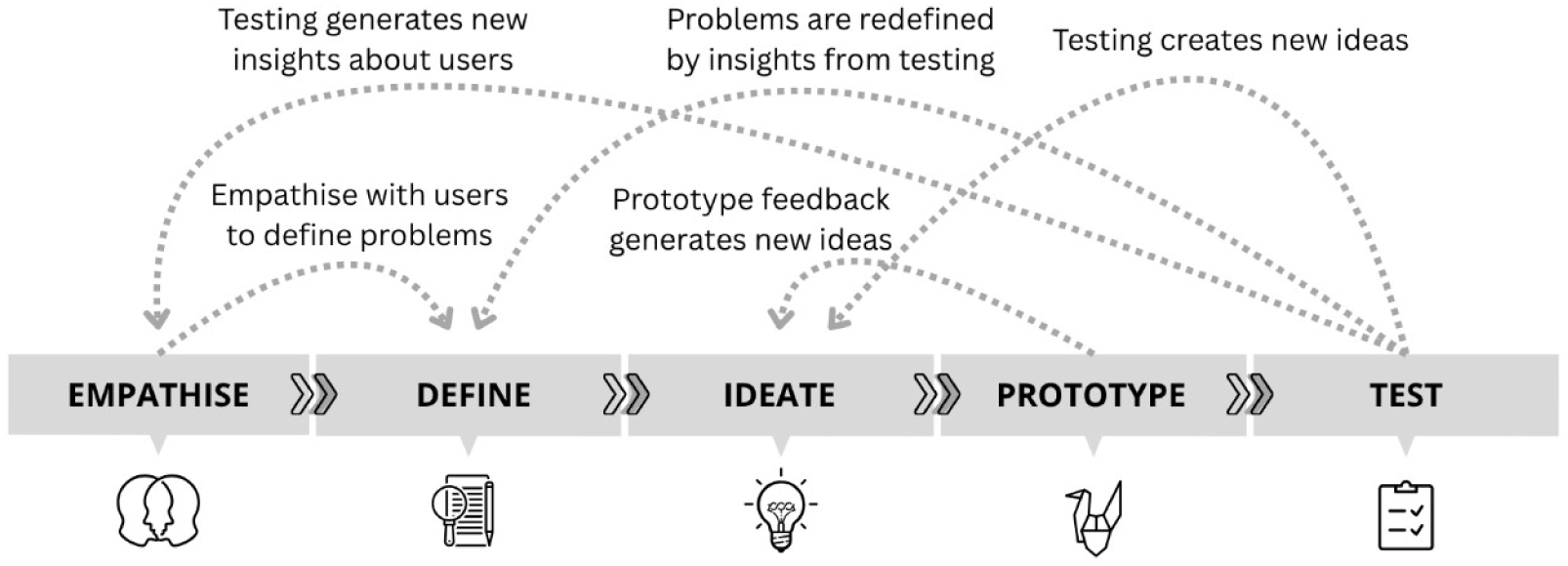
The Non-Linear Nature of the Five-Stage Design Thinking Framework. Note. Adapted from ‘The 5 Stages of Design Thinking’ by the Interaction Design Foundation (CC BY-NC-SA 3.0).

While the use of user- and human-centred design frameworks is often cited in the digital mental health literature as a requirement for designing effective and engaging digital tools (Berardi et al., 2024; Jacob et al., 2020), there is growing consensus on the limitations of user- and human-centred design (Forlizzi, 2018), in particular in the eHealth domain (Van Velsen et al., 2022).

Healthcare service-delivery infrastructure is complex, and typical human-centred design frameworks may not adequately account for the wider scope of stakeholders, organisational systems, and service-delivery contexts (Altman et al., 2018; Eriksson et al., 2024; Forlizzi, 2018; Oliveira et al., 2021; Van Velsen et al., 2022). More holistic context-sensitive frameworks are better suited for the complexities of designing for healthcare environments (Kip et al., 2025) but there are relatively few successful case studies showcasing an end-to-end design process (Adler et al., 2025; Oliveira et al., 2021). To address these limitations, this project expands the traditional Design Thinking framework to facilitate a contextually sensitive, ’more-than-human’ approach. Rather than framing the human user as an isolated agent, we define our more-than-human lens as one that views human participants as inseparable components of a complex, relational sociotechnical environment (Coskun et al., 2022; Wakkary, 2021).

Within the complexity of a healthcare environment, this means that human behaviours are viewed as fundamentally entangled with an array of non-human systemic actors, including software infrastructures, diagnostic tools, physical clinic layouts, and organisational systems and processes (Coskun et al., 2022; Wakkary, 2021). Adopting a more-than-human design framework prompts the designer to treat individual user feedback not as isolated data, but as just one interdependent element of a larger, sociotechnical usage context. Design decisions are therefore expanded to account for the structural constraints, physical environments, and organisational realities in which human actors such as practitioners, patients, and other healthcare stakeholders operate.

To operationalise this, we introduced an intermediate ’Contextual Fit’ phase into the five-stage Design Thinking cycle (*see Figure 2*), enhancing the designers’ ability to navigate the sociotechnical complexity of healthcare settings.

**Figure 2.**
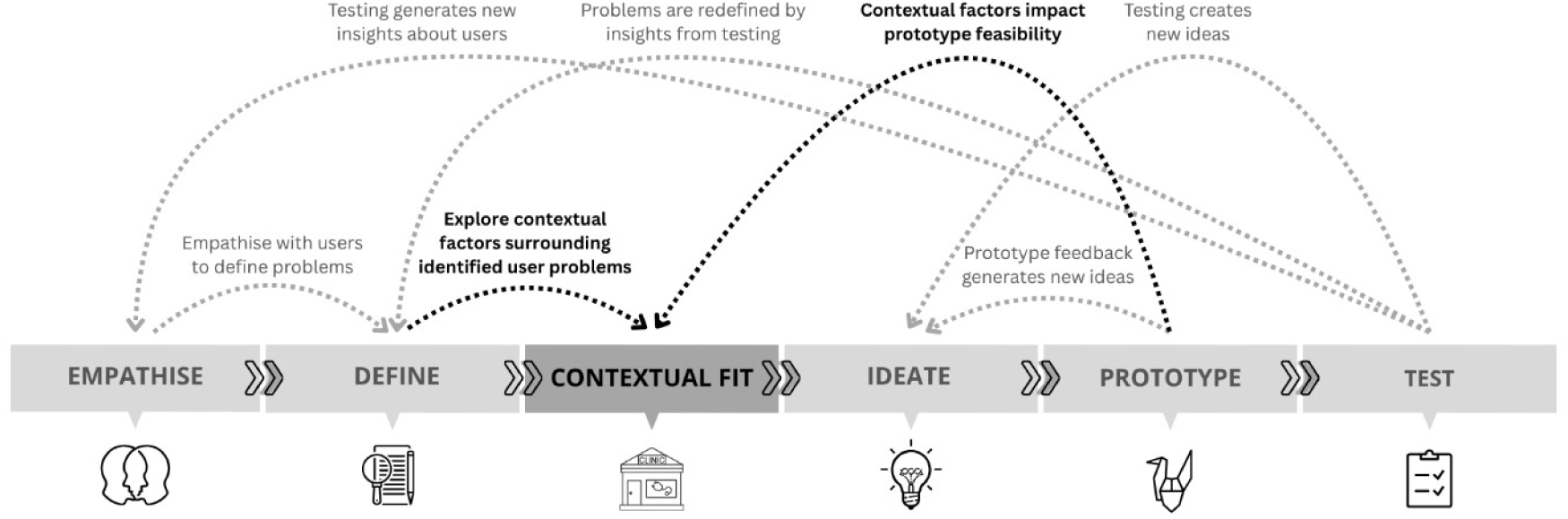
The Five-Stage Design Thinking Framework modified with an additional ‘contextual fit’ stage. Note. Adapted from ‘The 5 Stages of Design Thinking’ by the Interaction Design Foundation (CC BY-NC-SA 3.0).

### 1.5 Research Objectives

This paper details the design process of a DMHT to improve the scalability and effectiveness of an integrated primary care service in New Zealand’s public health system. The specific objectives of this research were:

1. To demonstrate the application of a context-sensitive more-than-human design framework to the development of a contextually relevant DMHT that serves the needs of practitioners, their patients, and functions effectively within a complex primary healthcare environment.
2. To provide a transparent, step-by-step case study of the iterative ideation and prototyping stages for one specific software feature, demonstrating the usefulness of design tools such as usage-context storyboards for navigating the complexities of the primary care service-delivery context.

## 2. Case Study Methods

### 2.1 Understanding the Context and Stakeholders

The project began with the Empathise stage of the Design Thinking framework: an exploration of the IPMHA model and the New Zealand primary care service-delivery context to discover how the model was being delivered and whether there was scope for improving service scalability. After identifying several IPMHA service-delivery challenges and opportunities for digital innovation, we then engaged with HIPs to understand their experiences working within the model. This was accomplished through interviews and a workforce survey, as well as ongoing engagement with various IPMHA organisational stakeholders to better understand the contextual factors surrounding HIPs’ reported experiences. This stakeholder engagement was vital for understanding the broader systemic and contextual variables influencing how HIPs work, providing a more holistic picture of the problem space. Through this process a variety of service-delivery challenges faced by HIPs were identified, many of which were well suited to be addressed by a purpose-built digital tool.

We combined the direct feedback from HIPs with the contextual insights gathered from engagement with stakeholders within the wider IPMHA ecosystem, synthesising this data into a set of user stories (Lucassen et al., 2016). Reflecting on the proposed usage context, we then refined these user stories into a collection of actionable problem statements to guide software feature ideation (Dam, 2025). These project resources acted as a touchstone for the design team, ensuring that the feature concepts were grounded not just in validated user problems, but also in the organisational and operational realities of the IPMHA service-delivery environment.

### 2.2 Concept Ideation and Iterative Prototyping

Once we had defined a set of problems to be addressed, we moved into the context-sensitive iterative ideation and prototyping cycle that is described in the following case study. Initial concept ideation involved collaborative sessions such as brainstorming and whiteboarding within our multidisciplinary design team. These concepts were then translated into low-fidelity prototypes, typically as pen-and-paper sketches. To guide the iterative refinement of our prototypes, we gathered qualitative feedback from HIPs through one-on-one, unstructured ‘think-aloud’ interviews (Boren, 2000). To enhance context-specificity and ground stakeholder feedback in real-world clinical scenarios, we presented the interface prototypes alongside usage-context storyboards.

Insights from each interview directly informed the next design iteration. This iterative cycle, incorporating both direct user feedback and contextual considerations, allowed for the progressive refinement of concepts and, when necessary, a return to the contextual fit and concept ideation stages to develop new solutions based on an updated understanding of user needs and contextual constraints. Prototypes evolved from low- to high-fidelity as the feature became more validated, culminating in a final design used to guide the development of a functional software minimum viable product (MVP).

### 2.3 Ideation and Prototyping Tools

During the design process a range of tools were used. Early ideation sessions relied on simple materials such as pen, paper, and whiteboards to facilitate collaborative brainstorming and the rapid generation of feature concepts. As ideas matured, they were translated into tangible prototypes for users to interact with. Initial low-fidelity prototypes were created as simple pen-and-paper sketches (Buxton, 2007). For more refined, higher-fidelity interactive prototypes, the team used the digital design tool Figma (Figma Inc., 2025). Finally, the validated high-fidelity Figma designs were used to guide the development of a functional MVP, which was built as a mobile Android app using the Flutter programming language (Google, 2025).

Usage-context drawings were used at all stages of the design and development process. These simple pen-and-paper sketches outlined the in-context usage of the software. They enabled a deeper understanding of the feedback received from stakeholders, clarified the function of contextual variables on service-delivery problems reported by HIPs and other IPMHA stakeholders, facilitated a shared understanding of usage-context during collaborative interdisciplinary design team activities, and aided in communicating functional requirements to the external software development team.

### 2.4 Participant Recruitment

A total of eight eligible participants from a single Primary Health Organisation (PHO) were recruited and successfully completed a series of iterative think-aloud (Boren, 2000) user feedback interviews. All participants were employed as HIPs in a New Zealand primary care clinic. Recruitment was facilitated through an electronic flyer distributed via industry contacts at the PHO with whom we were collaborating. Prospective participants received detailed information regarding the purpose of the study and its place within the broader digital tool development research project. Inclusion criteria required participants to be at least 18 years of age, currently employed in the HIP role at our collaborating PHO, and provide informed consent. To ensure confidentiality, all personally identifiable information was removed from the interview data. Informed consent was obtained for all participants. This study was approved by the Auckland Health Research Ethics Committee (reference number AH21908).

### 2.5 Data Collection

To gather iterative feedback on our prototypes we employed a qualitative research design using one-on-one interviews to explore HIPs’ perceptions regarding the usefulness and usability of the software tool prototype. Interviews were conducted as needed over a ten-month period, in alignment with the prototype development schedule. These unstructured interviews followed a think-aloud methodology (Boren, 2000), allowing participants to provide real-time feedback on the usability of the software features and share insights on how these features could integrate into their workflows and organisational systems. Early-stage prototypes were typically static pen-and-paper sketches, while later-stage prototypes were semi-functional click-through Figma (Figma Inc., 2025) wireframes.

This was a labour-intensive process that depended heavily on the significant time commitment of two key groups. Firstly, front-line HIPs provided crucial, detailed feedback on the prototypes as they evolved. Secondly, ongoing consultation with IPMHA organisational stakeholders helped our team to map the strategic landscape, clarifying both the opportunities for innovation and the practical limitations of integrating a new digital tool within the existing IPMHA service-delivery context.

### 2.6 Interview Structure

The interviews began with an introduction to the prototype and a description of the proposed usage context. This was supported by usage-context drawings showing the intended functionality of the software within the clinical environment. As participants interacted with the prototype, they were encouraged to ’think aloud’, sharing their expectations, experiences, and any difficulties they encountered. Throughout the process, the interviewer probed for clarification or feedback on particular features. The interviews were recorded and transcribed for analysis. After each interview, transcripts were analysed and used to refine subsequent prototypes, ensuring continuous development driven by user insights.

### 2.7 Data Analysis

Content analysis was chosen as the most appropriate analytical method for this data set, aligning with the study’s iterative design process and the deductive nature of the interviews (Krippendorff, 2019). This systematic framework was well-suited for the analysis of the unstructured think-aloud interview transcripts, allowing for actionable user feedback to be rapidly synthesised, directly informing the iterative refinement of subsequent prototypes in preparation for ongoing interview sessions. The analyses were conducted by the primary investigator using NVivo 12 software.

## 3. Case Study Results

Given the complexity of the proposed digital solution, a detailed process description covering the development of the entire software tool would be prohibitive. Therefore, this case study focuses on the development of a single feature within the overarching software package to illustrate the practical application of our context-sensitive adaptation of the Design Thinking framework. The feature that was showcased for this case study was the digital optimisation of the delivery, analysis, and recording of psychometric measures. As detailed in the subsequent process description (*see Figure 3*), our initial feature concept was a poor fit for the IPMHA service-delivery context. This mismatch required a return to the contextual fit and ideate stages with an updated understanding of user needs and usage-context to develop a more suitable solution. This iterative cycle demonstrates a core strength of a context-sensitive design framework: its capacity to ensure that design solutions remain grounded in the practical constraints of the service-delivery context as well as the needs of multiple stakeholders.

**Figure 3.**
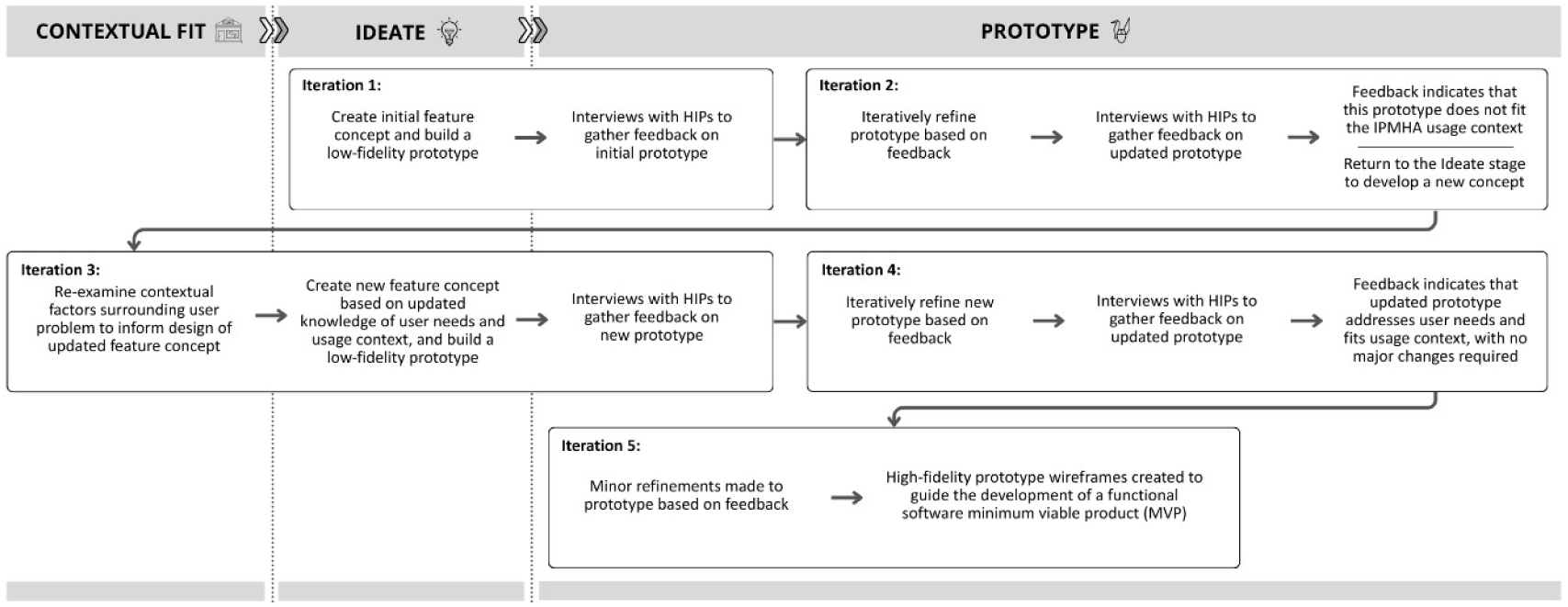
Overview of Design Iterations During the Ideation and Prototyping Stages for the Development of the Psychometric Digital Optimisation Feature.

### 3.1 Iteration 1: Initial Design Concept for Pre-Session Delivery of Psychometrics

#### 3.1.1 Concept and Design

Through a series of ideation sessions, including brainstorming, whiteboarding, and role-playing, our design team explored ideas for how this particular software feature could function. The team included members from a variety of different academic and applied backgrounds, including psychological medicine, human computer interaction, applied behaviour analysis, software engineering, and user-experience design.

From these ideation sessions, the concept that was carried forward into the prototyping stage was a software tool that allowed for a pre-session psychometric assessment to be completed by the client in the clinic waiting area (*see Figure 4*). In addition to streamlining psychometric data collection, analysis, and recording, this pre-session period could also be leveraged to provide clients with information about the HIP and what to expect in the upcoming session. This could help the client to feel better prepared when meeting their HIP for the first time, and provide an opportunity to pre-emptively address any misapprehensions about the content of the fACT session.

**Figure 4.**
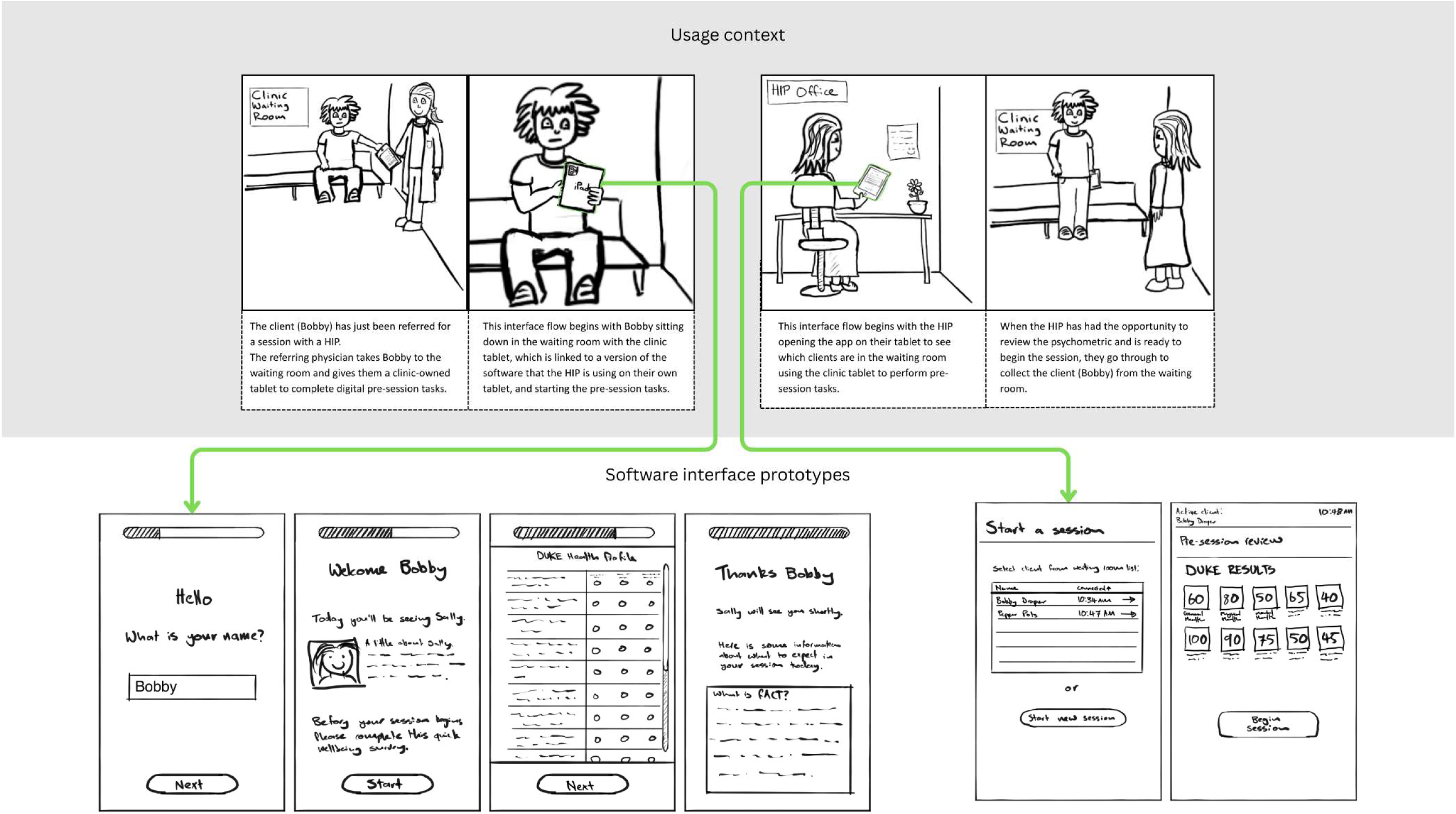
Storyboard Showing the Usage Context and Low-Fidelity Interface Drawings for Iteration 1.

A key motivation for using the waiting room downtime was the reported infrequency of the ‘warm handover’ from the referring physician. Ideally, the referring physician would directly introduce the client to the HIP, providing a verbal endorsement to establish rapport and initiate the therapeutic alliance. However, in practice, clients were frequently left unaccompanied in the waiting area, with no ‘warm handover’ from the physician. This context-specific service-delivery challenge highlighted an opportunity to capitalise on this waiting room time by engaging clients in a structured pre-session activity. With the digital delivery of a psychometric assessment, the proposed solution would not only enhance the efficiency of psychometric data collection but could also provide clients with a purposeful task while they waited to see the HIP. The proposed solution was a software tool on a clinic-provided tablet for completing a pre-session psychometric measure.

The software interface prototypes in Figures 4 and 5 are made up of basic pen-and-paper sketches showing the functionality of the software concept. While they do not look very refined or ‘professional’, this was an intentional design choice. The primary purpose of these pen-and-paper interface drawings was to serve as a prompt for gathering user feedback on functionality and usefulness. The use of low-fidelity sketches is a common strategy used in user-experience design and software development (Buxton, 2007). Keeping the design simple helps in two ways: it directs users’ focus toward functionality rather than design aesthetics and reinforces the perception that the prototypes are still in development. This, in turn, encourages users to provide candid, critical feedback on aspects they find ineffective or unhelpful.

**Figure 5.**
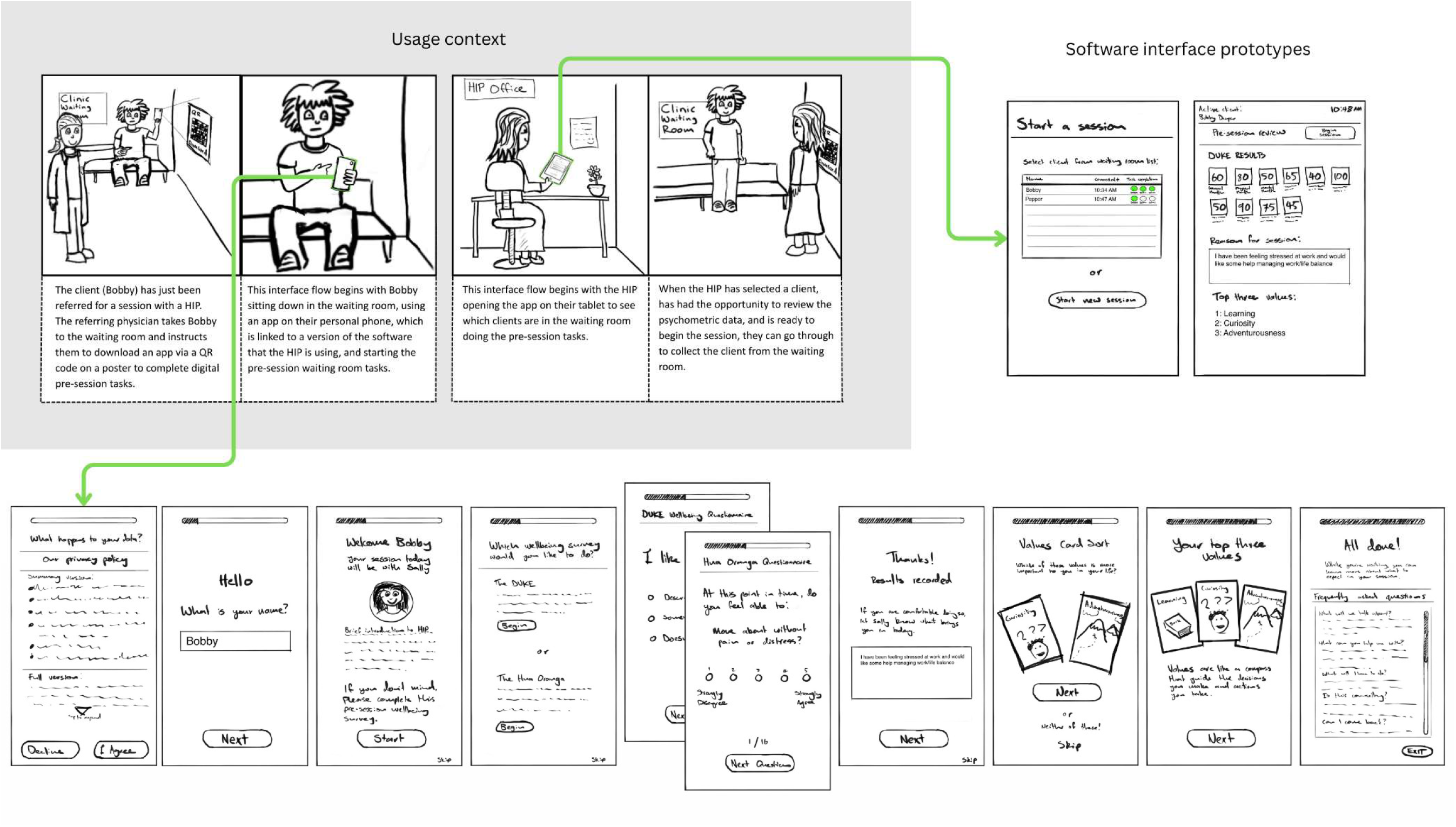
Storyboard Showing the Usage Context and Low-Fidelity Interface Drawings for Iteration 2.

#### 3.1.2 Feedback Summary: Concerns About Device Security and Systems Integration

Feedback on the initial prototype was largely positive regarding its core purpose. HIPs expressed a clear appreciation for the potential time-saving benefits, noting that having psychometric assessments completed and scored prior to the appointment would be particularly useful as they could then use the outcome data as a prompt during the contextual interview portion of the fACT session.

However, the feedback also raised numerous important logistical and practical questions that revealed the complexity of implementation in a primary care clinical setting. On a logistical level, there was significant uncertainty about who would be responsible for purchasing and managing the clinic devices: the clinic owner, the individual HIP, or the PHO. This was linked to concerns about the physical security of these devices and how to prevent theft. From a technical standpoint, practitioners questioned whether the software would integrate with existing patient management software (PMS) to save data to a client’s electronic health record, and if so, how. They also needed a system to notify them when a client had completed their pre-session tasks.

The client experience was a major focus. Feedback strongly emphasised that using the digital device must be optional to avoid disadvantaging clients who may not be comfortable with the technology. This led to questions about data privacy, how clients would be informed about the use of their data, and how informed consent would be obtained. Several content suggestions were made, such as adding a field for clients to state their primary concern and offering a choice of assessment tools beyond the Duke, like the Māori-focused Hua Oranga wellbeing measure. Despite these challenges, we determined the feature had enough potential to warrant a second iteration that would attempt to solve these practical issues.

### 3.2 Iteration 2: Shifting to a ’Bring Your Own Device’ Approach

#### 3.2.1 Concept and Design

Feedback from the first iteration made it clear that a clinic-provided device was logistically problematic. Therefore, the second iteration was redesigned around a ’bring your own device’ model, where clients would use their own smartphones to complete the pre-session psychometric. The proposed workflow involved a poster in the clinic with a QR code for the client to join the clinic WiFi and download the app. An alphanumeric or QR code would then link the client’s app to the profile of the HIP in that clinic, allowing the practitioner to see who was in the waiting room and monitor their task completion.

This refined prototype (*see Figure 5*), still rendered in low-fidelity drawings to encourage feedback, incorporated previous content suggestions. It included a way for the client to state what they needed help with, a values-elicitation task (a card sort) to complete pre-session, and replaced the block of text about the fACT model with a more engaging FAQ section. To address privacy concerns, a summarised privacy policy was included, along with clear descriptions for the choice of wellbeing measures.

#### 3.2.2 Feedback Summary: The Pre-Session Waiting Room Concept is Not Viable

While practitioners appreciated the inclusion of the privacy policy, the choice of measures, and the opportunity for clients to state their needs, the feedback on the overall workflow remained mostly negative. Significant concerns were raised about the feasibility of implementing these tasks in a busy primary care clinic. The administrative burden placed on the client: downloading an app, linking their account, reading agreements, was considered too time-consuming and a significant technology barrier, especially for clients who may already be feeling overwhelmed. It was noted that even with their own devices, clients would likely still require instruction from clinic staff, which we knew based on input from clinic stakeholders was not likely to happen.

Furthermore, the practicality of the pre-session workflow was questioned. The variable nature of clinic wait-times and the continued (though infrequent) practice of “warm handovers” would often not leave enough time for consistent use of the software. The values card sort, while a nice feature, was identified as being too time-consuming for the waiting room. A critical point was repeatedly emphasised: the need for this entire process to be strictly optional, as there was a strong concern it could be pushed by clinic staff as a prerequisite, thereby disadvantaging clients unable or unwilling to use it. The user feedback and wider contextual exploration made it clear that while some features were valuable, the entire pre-session concept was not appropriate for implementation into a busy primary care clinic. This led to the decision to abandon the waiting room concept and go back to the contextual fit and ideation stages to come up with a solution that was better suited to the service-delivery context. This was not a failure of our design, but a success of the context-sensitive iterative process that allowed the design team to identify critical flaws in the early stages of development.

### 3.3 Iteration 3: The Pivot - Return to the Ideation Stage to Design an In-Session Solution

#### 3.3.1 Concept and Design

The feedback for iteration two demonstrated that a pre-session solution was not viable for the primary care service-delivery context. This required a return to the ideation and contextual-fit stages to explore alternative strategies for reducing the time required for psychometric administration. Informed by contextual insights from clinic stakeholders and previous iterative interviews, subsequent ideation sessions led to a reorientation of our design strategy, discarding the waiting room concept to focus on improving the efficiency of in-session psychometric administration. The benefits of a more-than-human design framework were becoming increasingly evident at this stage. By prioritising broader contextual insights over the needs of one specific user, we were able to reorient our focus from the client to the practitioner as the primary software user for this feature. The new goal was to create a practitioner-facing tool to improve the efficiency of in-session psychometric administration: digitising the delivery of psychometric assessments (e.g., Duke and Hua Oranga) and automating scoring. This would reduce the in-session time required while still enabling HIPs to use the immediate results as prompts during the contextual interview. This feature development did not happen in isolation; the new prototypes (*see Figure 6*) were embedded into other features that were already at a more advanced stage of development. This allowed us to incorporate the new psychometric feature prototypes into more established user interface designs.

**Figure 6.**
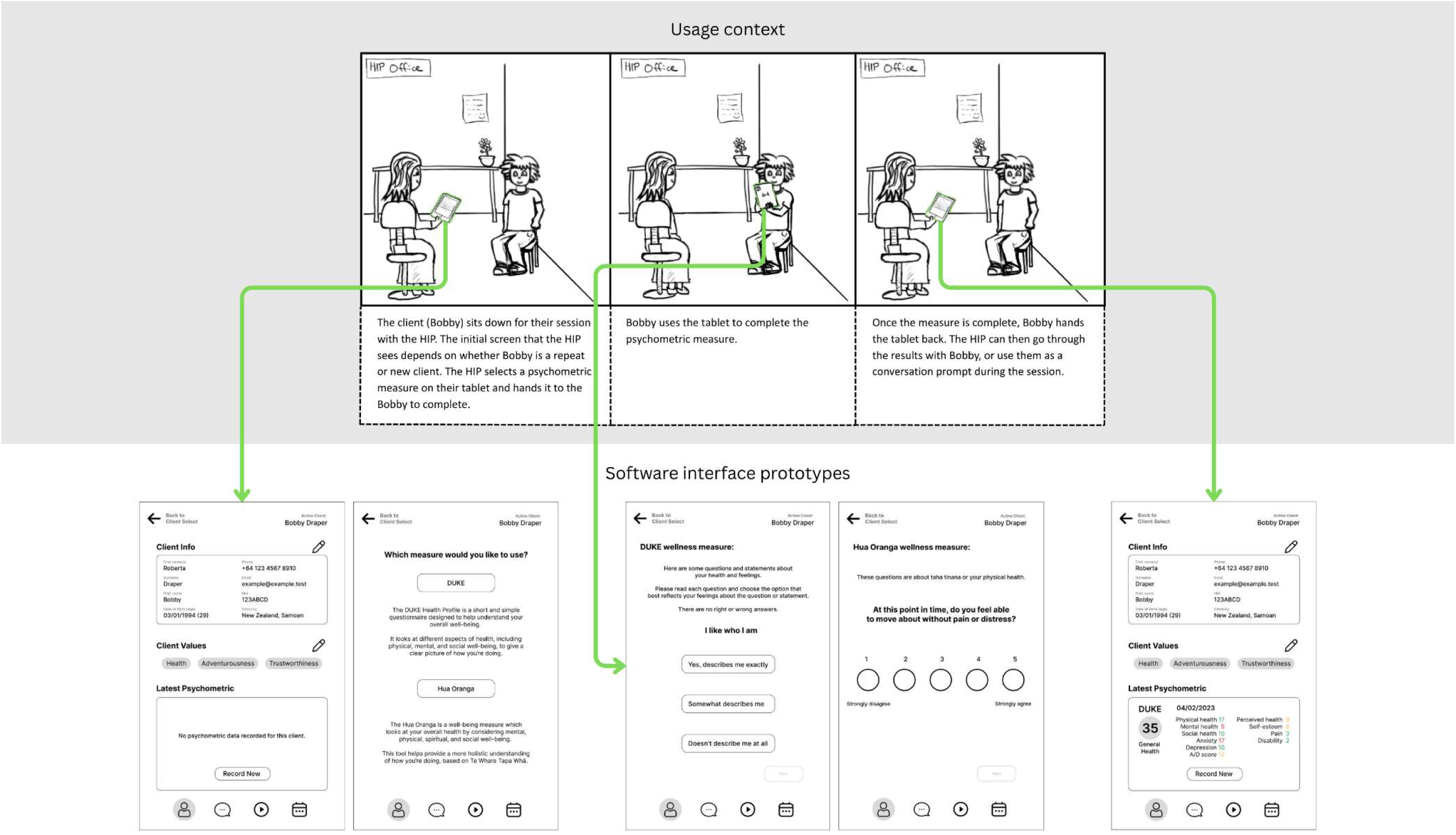
Storyboard Showing the Usage Context and Low-Fidelity Interface Drawings for Iteration 3.

#### 3.3.2 Feedback Summary: A Promising Concept With Untapped Potential

The new prototypes were met with positive feedback. HIPs confirmed their viability and began suggesting expansions to the functionality of the features. Because HIPs often need to refer clients to other services with their own screening requirements, it was suggested that the software include digital versions of additional measures such as the PHQ-9 and Kessler-10 scales (Kroenke et al., 2001; Kessler et al., 2002), to streamline referral processes for these additional stakeholders. To protect sensitive information, HIPs suggested that the software should require a security pin to change screens when a client uses a practitioner’s tablet to prevent the client from navigating to other parts of the software.

Feedback also highlighted a desire for better data visualisation. A feature that would allow practitioners to view a client’s entire psychometric history in a single, consolidated view was described as “fantastic.” This would eliminate the need to search through past clinical notes and provide the ability to compare different measures and identify trends at a glance. Finally, a suggestion was made to extend the tool’s use beyond the in-clinic session; if clients could access a wellness measure on their own device after their appointment, it would provide a valuable method for gathering post-session updates and outcome data. This feedback validated the in-session approach and provided a clear direction for feature improvement and expansion.

### 3.4 Iteration 4: Refining the In-Session Tool With Post-Session Functionality

#### 3.4.1 Concept and Design

At this stage, we had a clear understanding of the needs of the various stakeholders, and the constraints of the primary care service-delivery context, allowing us to move to a medium-fidelity prototype with a more refined user interface. Iteration four focused on integrating the features requested in the previous round. We added additional psychometrics: PHQ-9, Kessler-10, and SDQ (Kroenke et al., 2001; Kessler et al., 2002; Goodman, 1997) for HIPs to make referrals, incorporated a security check to restrict clients from navigating away from the questionnaire screen, and created a dedicated screen for HIPs to view a client’s entire psychometric history.

The most significant design change was the integration of post-session psychometric collection. This was made possible by a client-facing companion app that was being developed in parallel. The prototype was designed so that clients could track their wellbeing on their own device after a session, with ongoing outcome measure results being transmitted back to the HIP for remote progress monitoring. The functionality was illustrated in updated storyboards showing both the in-session practitioner workflow (*see Figure 7*) and the post-session client workflow (*see Figure 8*).

**Figure 7.**
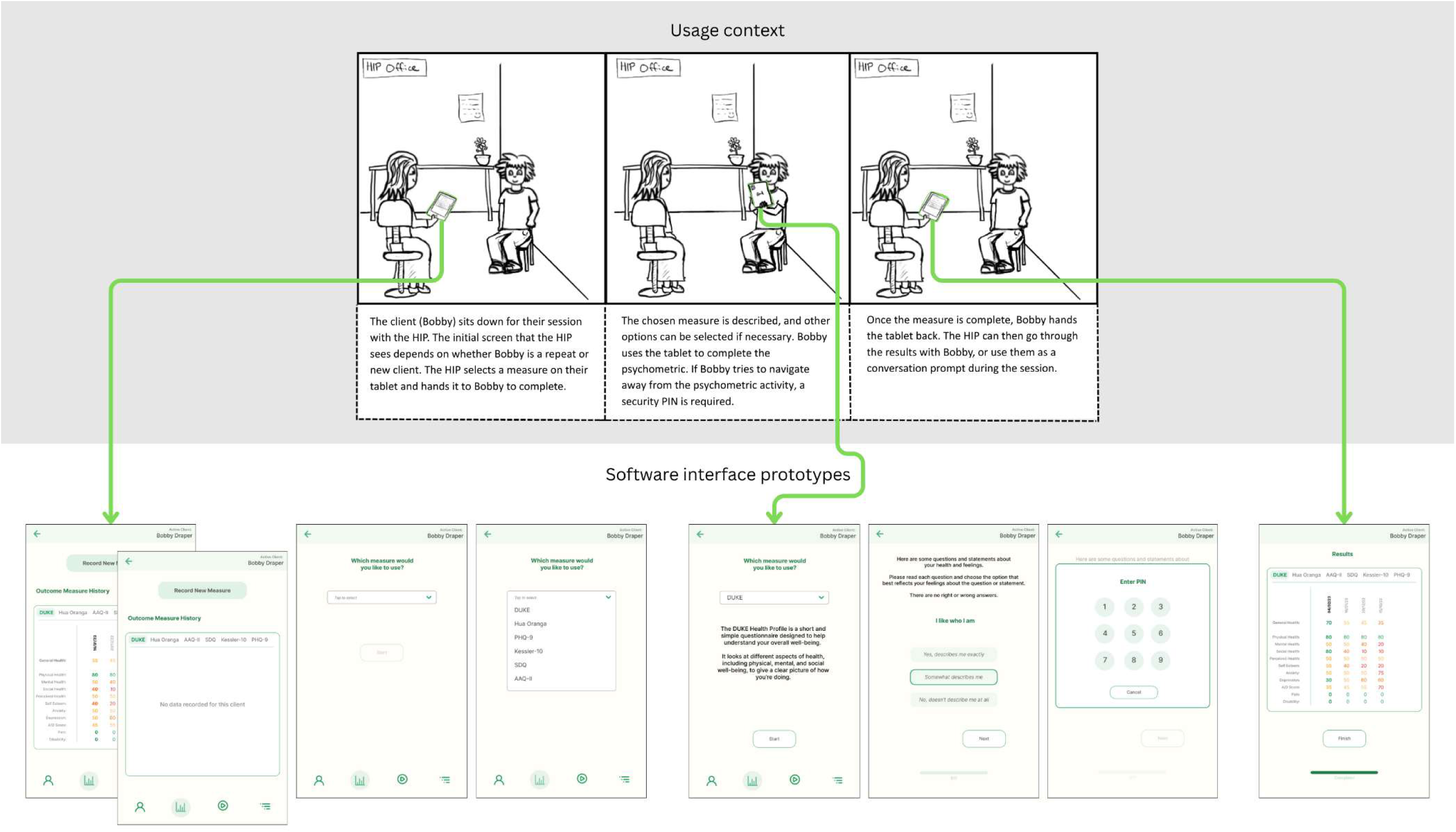
Storyboard Showing the Usage Context and Interface Designs for Iteration 4 (In-Session).

**Figure 8.**
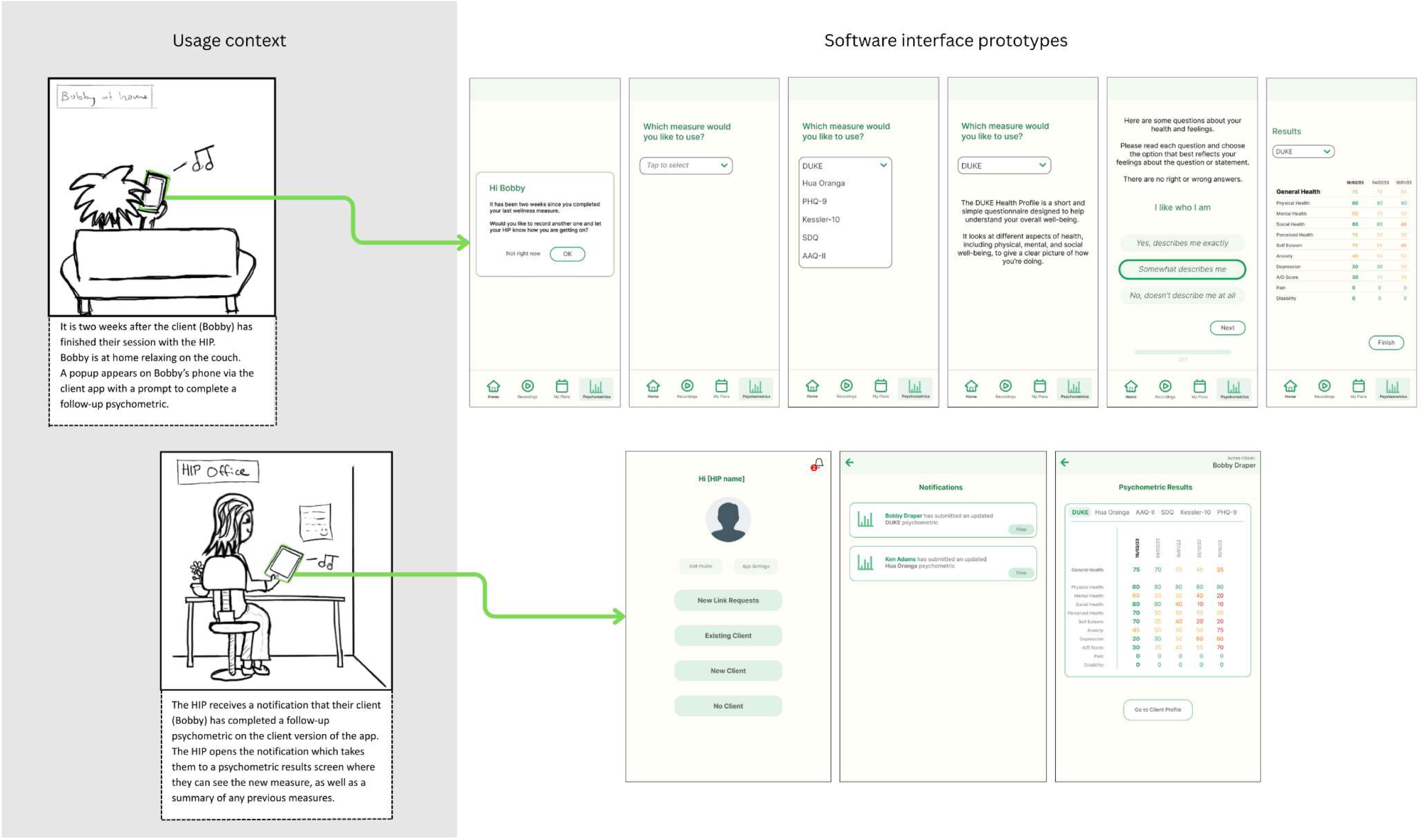
Storyboard Showing the Usage Context and Interface Designs for Iteration 4 (Post-Session).

#### 3.4.2 Feedback Summary: Prototype Fits Workflow and Clinic Context, Minor Improvements Requested

Feedback confirmed that the prototype had the potential to significantly reduce the administrative burden of psychometric delivery. The ability to use the results immediately in-session to guide the contextual interview was repeatedly mentioned as highly desirable. The integration with the client companion app for automated post-session follow-up was viewed very positively, not only as a tool for improving clinical practice but also as a way for practitioners to see the impact of their work. It was suggested that to improve client engagement, the client should be made explicitly aware that these follow-up measures serve as progress updates for their HIP, reinforcing the social contract and extending the benefits of the therapeutic alliance.

Several practical recommendations were made for improving implementation into HIP workflows. To be more efficient, when a practitioner taps a notification about a client update, it should take them directly to the client’s main profile page. To ensure the system is manageable, HIPs stressed the need to limit the frequency of psychometric updates to avoid being overwhelmed. Clear boundaries were also requested, limiting post-session access to general wellness measures like the Duke or Hua Oranga, while restricting clinical referral tools like the PHQ-9 to in-session use only.

### 3.5 Iteration 5: Final High-Fidelity Prototype to Guide MVP Development

#### 3.5.1 Concept and Design

At this stage, feedback included only minor refinements, indicating that the core functionality was complete and well-validated. Perfect is often the enemy of progress, so at this stage we felt it was time to wrap up the development cycle for this feature. Based on the feedback for iteration four, we made final functional adjustments and refined the interface to ensure consistency with other features in the software package.

The specific changes included adding text to the client companion app to remind the client that their wellness measure results will be sent to their HIP. The client-side app interface was modified with a dark green colour palette and a fern motif to appear more inviting and less ‘clinical’. Client-initiated psychometrics were limited to the Duke or Hua Oranga, defaulting to whichever one they used in-session. Finally, a system was designed where reminders to complete a measure would only be shown to clients every two weeks. These changes were incorporated into high-fidelity prototypes that reflected what the final software would look like, illustrated in detailed storyboards for both in-session (*see Figure 9*) and post-session (*see Figure 10*) functionality.

**Figure 9.**
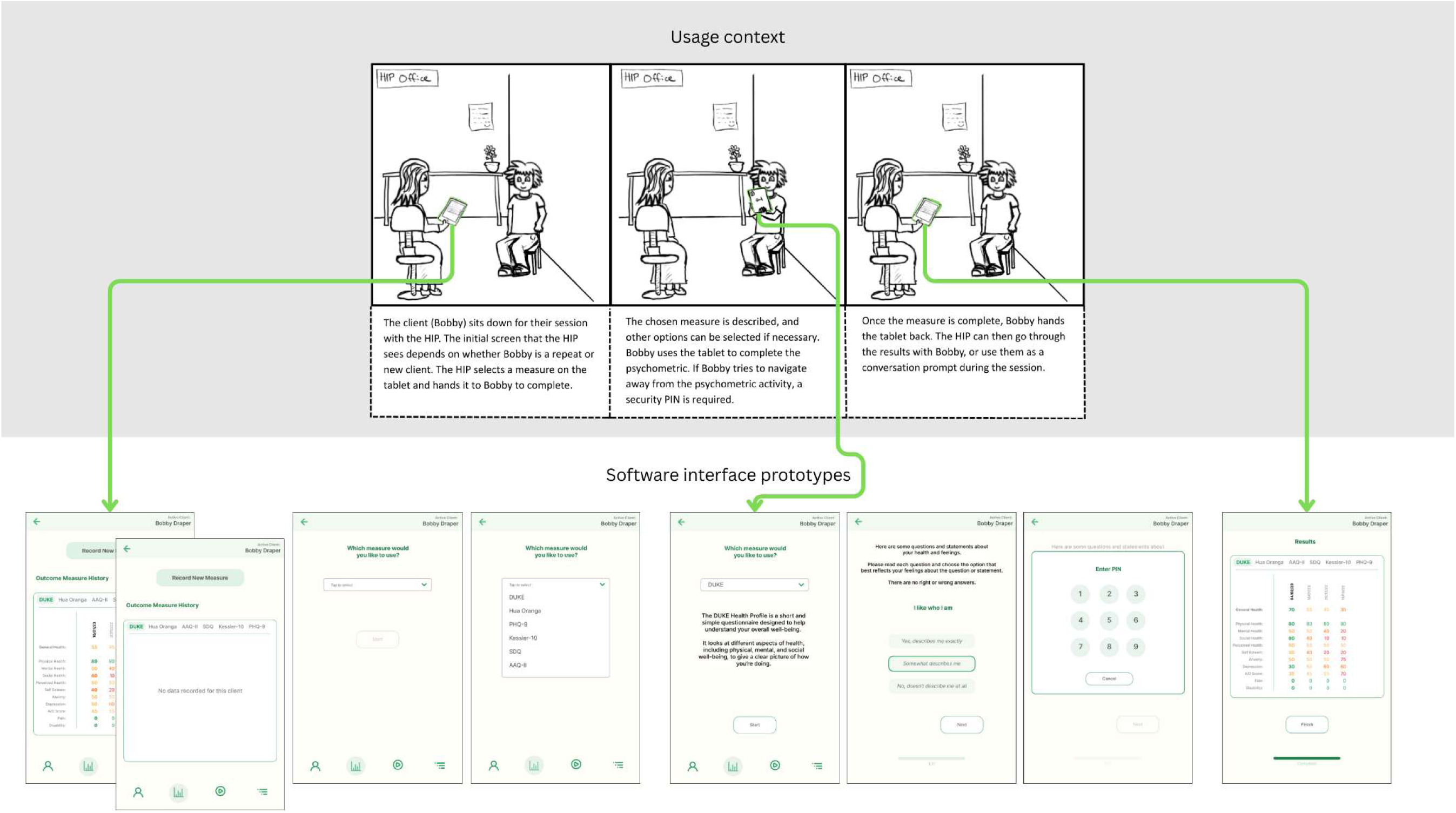
Storyboard Showing the Usage Context and Interface Designs for Iteration 5, the Final High-Fidelity Prototype (In-Session).

**Figure 10.**
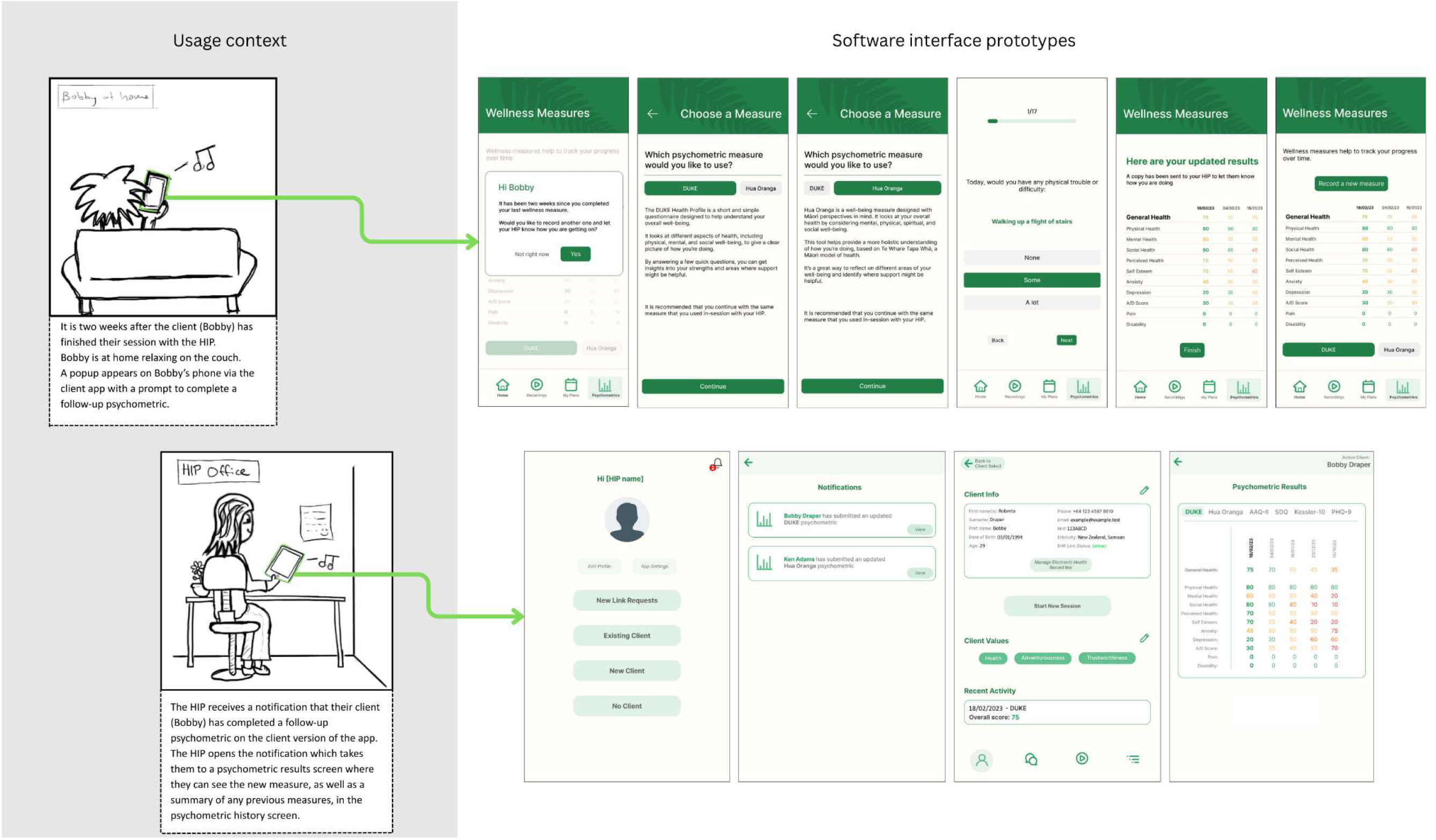
Storyboard Showing the Usage Context and Interface Designs for Iteration 5, the Final High-Fidelity Prototype (Post-Session).

#### 3.5.2 Feedback Summary: Requirements Met and Design Finalised

With minimal changes from the previous prototype, feedback for this final version confirmed that the software feature was well-suited for integration into the HIPs’ workflow and the IPMHA service-delivery context. No major usability issues were identified.

The feedback included two final refinements for data management. First, the process of automatically saving client-sent data should include a manual approval step by the HIP to ensure its validity. Second, a time limit should be implemented to eventually terminate the app link with a client, preventing indefinite updates. With these final checks, we were confident all core functional requirements had been accounted for. The next step was to use these high-fidelity prototypes to create detailed functional wireframes, which were then handed off to the software engineering team to inform the development of a functional software MVP.

## 4. Discussion

### 4.1 Principal Findings

This paper detailed the iterative design process of a digital mental health tool (DMHT) intended to support Health Improvement Practitioners (HIPs) within New Zealand’s Integrated Primary Mental Health and Addictions (IPMHA) model. We adopted a ’more-than-human’ design framework by applying the five-stage Design Thinking model (Brown, 2008, 2009) through a more contextually sensitive lens, ensuring that the complexities of the healthcare service-delivery context were considered alongside direct user needs. The framework’s value was proven by an early failure: our initial concept of a waiting-room tool, while user-friendly, was shown to be unworkable in the broader clinical environment. The pivot to a more contextually appropriate solution underscores the central argument of this paper: for DMHTs to be successfully implemented into complex environments like healthcare, a design focus that extends beyond one specific user to the wider usage context is essential. The development of this psychometric optimisation feature serves as a compelling case study showing the importance of adopting a context-sensitive more-than-human framework to account for the complexity of the healthcare environment (Eriksson et al., 2024; Oliveira et al., 2021).

### 4.2 Reflections on the Iterative Prototyping Process

A key takeaway from this project was the importance of maintaining a dual focus on the various humans engaging with the tool as well as the wider usage context. While user- and human-centred design is rightly cited as a prerequisite for creating effective digital tools (Berardi et al., 2024; Jacob et al., 2020), our experience aligns with a growing consensus that it can be insufficient in complex sociotechnical systems like healthcare (Eriksson et al., 2024; Forlizzi, 2018; Van Velsen et al., 2022; Oliveira et al., 2021). The failure of our initial waiting-room concept illustrates this limitation. The concept directly addressed a practitioner-stated need to save in-session time. However, iterative prototyping informed by contextual considerations revealed numerous environmental variables impacting the viability of our proposed solution. These included: inconsistent wait times, the lack of staff to manage devices, the administrative burden on distressed clients, and workflow disruptions. Shifting our perspective to encompass not just the practitioner and client, but also the surrounding clinical ecosystem, was essential. This wider, context-aware view revealed why the initial solution failed and enabled us to pivot toward a more appropriate design.

This process highlighted a limitation of design frameworks that revolve around the needs of one particular stakeholder (Eriksson et al., 2024). Our initial design concepts were predominantly built around the client as the primary stakeholder, focusing on how they would use the software in the waiting room. However, as our understanding of the primary care context grew, our designs evolved, and between iterations two and three we made a significant pivot from a focus on the client’s needs to those of the practitioner. This shift was driven by the design team gaining a deeper understanding of what was possible within this service-delivery context. Focusing primarily on the perspective of a single stakeholder may have caused us to overlook important contextual cues and potentially miss a necessary shift in focus to a different stakeholder.

The addition of an explicit ‘Contextual Fit’ stage prompted the design team to look beyond surface-level feedback and explore the function of observed and reported behaviours, identifying underlying environmental variables within the clinical ecosystem. This approach ensured that the final design was grounded in the contextual realities of the clinic’s organisational systems and processes.

This process also demonstrated that feature requirements can, and often should, evolve significantly during development, even after a thorough initial analysis of user needs. The pivot from a pre-session to an in-session tool was not a failure of our early-stage user-needs research but rather a success of the contextually sensitive iterative prototyping process. It showed that true usability and contextual fit emerge from an ongoing dialogue with a range of stakeholders and a deepening understanding of their environment throughout the entire development cycle (Coskun et al., 2022; Eriksson et al., 2024; Mohr et al., 2025).

However, this iterative process was not without its challenges. It was highly labour- and resource-intensive, requiring sustained cooperation among multiple stakeholders, including clinicians, patients, and organisational leaders. Conducting this type of agile research presented unique difficulties, particularly when navigating inflexible ethics approval processes that are often ill-suited to the rapid, responsive nature of iterative design.

### 4.3 The Role of Usage-Context Storyboards in Facilitating Effective Context-Centred Design

To effectively navigate the complexities of the healthcare environment, our team relied heavily on usage-context storyboards alongside interface prototypes. These storyboards proved to be an invaluable tool, serving as more than just a prompt for gathering feedback from practitioners. They also functioned as an interdisciplinary translation tool, creating a shared contextual understanding that bridged the communication gaps between the multidisciplinary design team and clinical stakeholders. For HIPs, the storyboards grounded abstract software concepts in familiar, real-world clinical scenarios, enabling more specific and relevant feedback. For the design and development teams, they maintained a consistent, shared vision of the usage context. They were also instrumental in communicating functional requirements to the software engineering team, providing an overview of the proposed functionality of the software that is often lost in technical requirements documents. This demonstrates how simple, low-fidelity tools can be powerfully effective in facilitating a context-centred design process.

### 4.4 Strengths and Limitations

A primary strength of this study is its rigorous application of a more-than-human design methodology, allowing for a deep iterative exploration that moved beyond surface-level user needs to address the complex sociotechnical realities of the primary care environment.

The detailed, transparent case study of the pivot from a pre-session to an in-session tool provides a powerful, practical example of this methodology in action, demonstrating how to identify and overcome contextual barriers early in development. By documenting this process, this research helps to fill a notable gap in the literature, which calls for contextually sensitive ‘more-than-human’ design frameworks to overcome implementation barriers (Eriksson et al., 2024; Kip et al., 2025) but offers few detailed examples of how to apply these within complex environments like healthcare (Adler et al., 2025).

This study also shows the benefits of collaborative interdisciplinary research, with complementary methodologies and expertise from human-computer interaction, contextual behavioural science, and psychological medicine being combined to facilitate the successful application of this context-sensitive ‘more-than-human’ product development framework. Another key strength is the effective use of design artefacts, particularly usage-context storyboards, which proved invaluable as a translation tool for facilitating shared understanding across a multidisciplinary team and with clinical stakeholders.

However, the study also has several limitations. First, the findings are based on a small sample of eight HIPs recruited from a single Primary Health Organisation in New Zealand. While this enabled deep context-specific feedback, it limits the generalisability of the results, as the specific contextual challenges and user needs may differ in other regions or healthcare systems. Second, while this paper details the design and prototyping phases of development, it does not evaluate the real-world implementation or clinical effectiveness of the final functional software MVP. The outcomes presented relate to the success of the iterative design process, not the tool’s ultimate impact on practitioner efficiency, model fidelity, or patient outcomes. A significant limitation of this research is that it did not directly include patient feedback, relying instead on practitioner perspectives to understand the client experience. While this practitioner-centred approach was necessitated by practical constraints within the academic context, such as securing ethical approval for iterative prototyping research with clinical populations, it is a notable shortcoming. Future research should therefore prioritise gathering direct feedback from patients to validate the design of any client-facing features.

### 4.5 Contributions

This research makes several key contributions to the field of digital mental health and human-computer interaction. First, it offers a detailed, practical case study that demonstrates how to proactively design a DMHT to overcome known barriers to practitioner adoption, moving beyond the general recommendations in the literature for the use of user- and human-centred design frameworks. Second, it provides a proof of concept for a context-centred ‘more-than-human’ design approach, showing the importance of understanding organisational systems and clinical workflows when designing for complex healthcare contexts. Finally, this study highlights the utility of specific design artefacts, namely usage-context storyboards, as a powerful tool for facilitating shared understanding and communication across the design team, clinical and organisational stakeholders, and software developers.

### 4.6 Future Research

The next phase of this research is to evaluate the complete functional software MVP. We will begin with usability testing with a cohort of HIPs to assess the software’s contextual fit and the usefulness of its features within a simulated clinical context. Feedback from this testing will guide the development of a clinically implementable version of the MVP. Following this, we plan to conduct a real-world implementation trial. This trial will measure the tool’s impact on service-delivery metrics such as model fidelity, administrative efficiency, and patient outcomes, and will incorporate feedback directly from HIPs, their clients, and other IPMHA clinical and organisational stakeholders.

### 4.7 Conclusion

The field of human-computer interaction is moving toward ‘more-than-human’ design frameworks to meet the demands of designing for increasingly complex service-delivery environments (Eriksson et al., 2024). This article offers a case study describing the design of a digital tool within one such complex environment: New Zealand’s integrated care IPMHA service. Eight practitioners participated in iterative design sessions, where usage-context was explicitly defined alongside the presentation of interface prototypes. This prioritisation of context in design decisions allowed for flexibility regarding the needs of additional stakeholders and contextual considerations, and led to a substantial pivot: the initial design had clients filling in psychometric data in the waiting room, but when more accurately accounting for the opportunities and barriers within the primary care clinic environment, the final design shifted that data entry to practitioners using the software in-session. This outcome shows how moving beyond a strictly user-centred design framework, incorporating the needs of multiple stakeholders and taking into account contextual variables, led to a better solution for use within a complex healthcare environment.

## Declarations

### Data Availability

The qualitative iterative feedback interview data are not publicly available to protect participant privacy. Aggregated data supporting the findings are available from the corresponding author upon reasonable request.

## Author Contributions

Vincent Allen (VA), Karolina Stasiak (KS), Danielle Lottridge (DL).

VA: Conceptualization, Project Administration, Investigation, Data Curation, Formal Analysis, Resources, Writing – Original Draft, Writing – Review and Editing.

KS: Conceptualization, Supervision, Writing – Review and Editing

DL: Conceptualization, Supervision, Writing – Review and Editing.

All authors have read and agreed to the published version of the manuscript.

